# Global Expansion of SARS-CoV-2 Variants of Concern: Dispersal Patterns and Influence of Air Travel

**DOI:** 10.1101/2022.11.22.22282629

**Authors:** Houriiyah Tegally, Eduan Wilkinson, Darren Martin, Monika Moir, Anderson Brito, Marta Giovanetti, Kamran Khan, Carmen Huber, Isaac I. Bogoch, James Emmanuel San, Joseph L.-H. Tsui, Jenicca Poongavanan, Joicymara S. Xavier, Darlan da S. Candido, Filipe Romero, Cheryl Baxter, Oliver G. Pybus, Richard Lessells, Nuno R. Faria, Moritz U.G. Kraemer, Tulio de Oliveira

**Author notes:** These authors contributed equally (Houriiyah Tegally, Eduan Wilkinson). These authors jointly supervised this work (Moritz U.G. Kraemer, Tulio de Oliveira).

## Abstract

In many regions of the world, the Alpha, Beta and Gamma SARS-CoV-2 Variants of Concern (VOCs) co-circulated during 2020-21 and fueled waves of infections. During 2021, these variants were almost completely displaced by the Delta variant, causing a third wave of infections worldwide. This phenomenon of global viral lineage displacement was observed again in late 2021, when the Omicron variant disseminated globally. In this study, we use phylogenetic and phylogeographic methods to reconstruct the dispersal patterns of SARS-CoV-2 VOCs worldwide. We find that the source-sink dynamics of SARS-CoV-2 varied substantially by VOC, and identify countries that acted as global hubs of variant dissemination, while other countries became regional contributors to the export of specific variants. We demonstrate a declining role of presumed origin countries of VOCs to their global dispersal: we estimate that India contributed <15% of all global exports of Delta to other countries and South Africa <1-2% of all global Omicron exports globally. We further estimate that >80 countries had received introductions of Omicron BA.1 100 days after its inferred date of emergence, compared to just over 25 countries for the Alpha variant. This increased speed of global dissemination was associated with a rebound in air travel volume prior to Omicron emergence in addition to the higher transmissibility of Omicron relative to Alpha. Our study highlights the importance of global and regional hubs in VOC dispersal, and the speed at which highly transmissible variants disseminate through these hubs, even before their detection and characterization through genomic surveillance.

**Highlights:**

- Global phylogenetic analysis reveals relationship between air travel and speed of dispersal of SARS-CoV-2 variants of concern (VOCs)
- Omicron VOC spread to 5x more countries within 100 days of its emergence compared to all other VOCs
- Onward transmission and dissemination of VOCs Delta and Omicron was primarily from secondary hubs rather than initial country of detection during a time of increased global air travel
- Analysis highlights highly connected countries identified as major global and regional exporters of VOCs

## Introduction

Since the emergence of SARS-CoV-2 in late 2019, multiple waves of infections have spread across the world. Successive waves have been caused typically by new variants, each of which replaced previously dominant variants due to higher transmissibility and/or ability to evade immunity. At the end of 2020, the first three variants of concern (VOCs), Alpha^1^, Beta^2^ and Gamma^3^ emerged. Along with some less successful novel lineages (termed variants of interest or VOIs), these VOCs were characterized by a combination of increased intrinsic transmissibility, sometimes enhanced immune evasion capabilities, and increased pathogenicity^4^. Each of the VOCs was associated initially with increasing SARS-CoV-2 incidence in their presumed countries of origin: Alpha in the United Kingdom^1,5^, Beta in South Africa^2^ and Gamma in Brazil^3^. In the second quarter of 2021, these three VOCs started being displaced throughout the world by Delta, a fourth VOC with substantially increased intrinsic transmissibility and pathogenicity compared to the initial three VOCs. Retrospective analysis revealed that, as with Alpha and Beta, Delta may have first arisen between September and October 2020^6^, but only spread globally after it caused a large outbreak in India in March 2021^7^. The most recent VOC, Omicron, was first detected during a rapid increase in cases in Botswana and South Africa in November 2021^8^.

At each successive stage of the pandemic, global transmission of SARS-CoV-2 has continued within a context of shifting public health responses, virus evolution, and dynamic changes in host immunity. The pandemic precipitated unprecedented changes in the intensity and nature of human mobility, both internationally, through strict restrictions on global travel, and nationally, via government-implemented public health and social measures (PHSM)^9^. While the intention behind initial restrictions on international travel were to limit the dispersal of the virus out of the first outbreak epicenters with the view to possible elimination, they were used later to attempt to limit or slow the dispersal of VOCs out of their perceived regions of origin and to reduce epidemic intensity. Following the global vaccination rollout, travel restrictions and other PHSM have been lifted gradually in most parts of the world, bringing travel and mobility patterns back towards similar levels seen prior to the pandemic^10^. Regardless of the intensity and range of travel restrictions, multiple SARS-CoV-2 variants have disseminated to, and risen to prominence across large swathes of the world.

Understanding the global dispersal patterns of SARS-CoV-2 VOCs in the context of local and global human mobility is critical if we are to evaluate objectively the relative importance of targeted travel restrictions as pandemic prevention and/or control measures. Fortunately, increased investments in genomic surveillance and data sharing throughout the pandemic have enabled the effective tracking of VOCs in near real-time, mainly through GISAID^11^. Consequently, sufficient genomic sequence data are now available to enable detailed investigations of past SARS-CoV-2 transmission dynamics in different locations and at different scales^6,12–17^. Yet, the factors underlying variability in the dissemination of SARS-CoV-2 VOCs are yet to be fully understood, especially on a global scale and when comparing all five VOCs.

Here, we combine phylogenetic models that leverage multiple sets of ∼20,000 genomes per VOC from >100 countries with global air passenger data to reconstruct the global spread of each VOC. We investigate the movement dynamics of each VOC at country and regional levels to determine source-sink dynamics and establish the regional and global contributions of individual countries to the exportation of VOCs. We specifically investigate the role of the countries that first reported an epidemic of each VOC on the global movement dynamics of VOCs and measure the influence of international travel on VOC dispersal.

## Results

### VOC Global Dissemination Patterns

To quantify the global dissemination patterns of each VOC, we performed ancestral state reconstruction of discrete spatial locations using dated phylogenetic trees that were inferred from a subset of representative sampled genomes (for which sequence sampling locations were known). Genomes were sampled in proportion to country and variant-specific case counts, and analyses were repeated across ten replicates of ∼20,000 randomly sampled genomes per VOC (after accounting for VOC specific case counts). Continental source-sink dynamics were determined by calculating the net difference between viral exportation and importation events for each country and continent (see Materials & Methods for more detail). A limiting factor of this analysis is that countries with under-reported incidence and low sequencing proportions^18^ but high global connectivity would have been missed as important global or regional VOC disseminators (given the reliance of our methods on genomic data and underlying testing patterns).

Our analyses reveal distinct global expansion processes for each VOC. The Alpha, Beta and Gamma variants co-circulated globally from November-December 2020 to June-July 2021 (Figure S1). As expected, Europe was a major source of the Alpha variant, with the United Kingdom (UK) contributing the highest estimated number of exports to the rest of the world (>2000; ∼50% of Alpha exports; Figure 1A). Global expansion can be described as a multi-stage process: first, at the end of 2020 and beginning of 2021, we estimate that Alpha spread mostly within Europe (>3000 exports between European regions), and from Europe to the Americas and Asia (>600 exports from Europe). Most introductions of Alpha to Africa were from Europe or North America (>60 exports), initially to West Africa then to East Africa (Figure 1A). Second, between February and May 2021, Alpha spread within the Americas and Asia, and we observe viral lineage exchanges from East Africa to Asia. We estimate that Africa and South America’s contributions to global dissemination of Alpha were minimal overall. It is important to note that due to subsampling and uneven sampling, viral importation numbers presented in this manuscript need to be interpreted as relative and are likely an underestimate of the actual number of importations.

**Figure 1:**
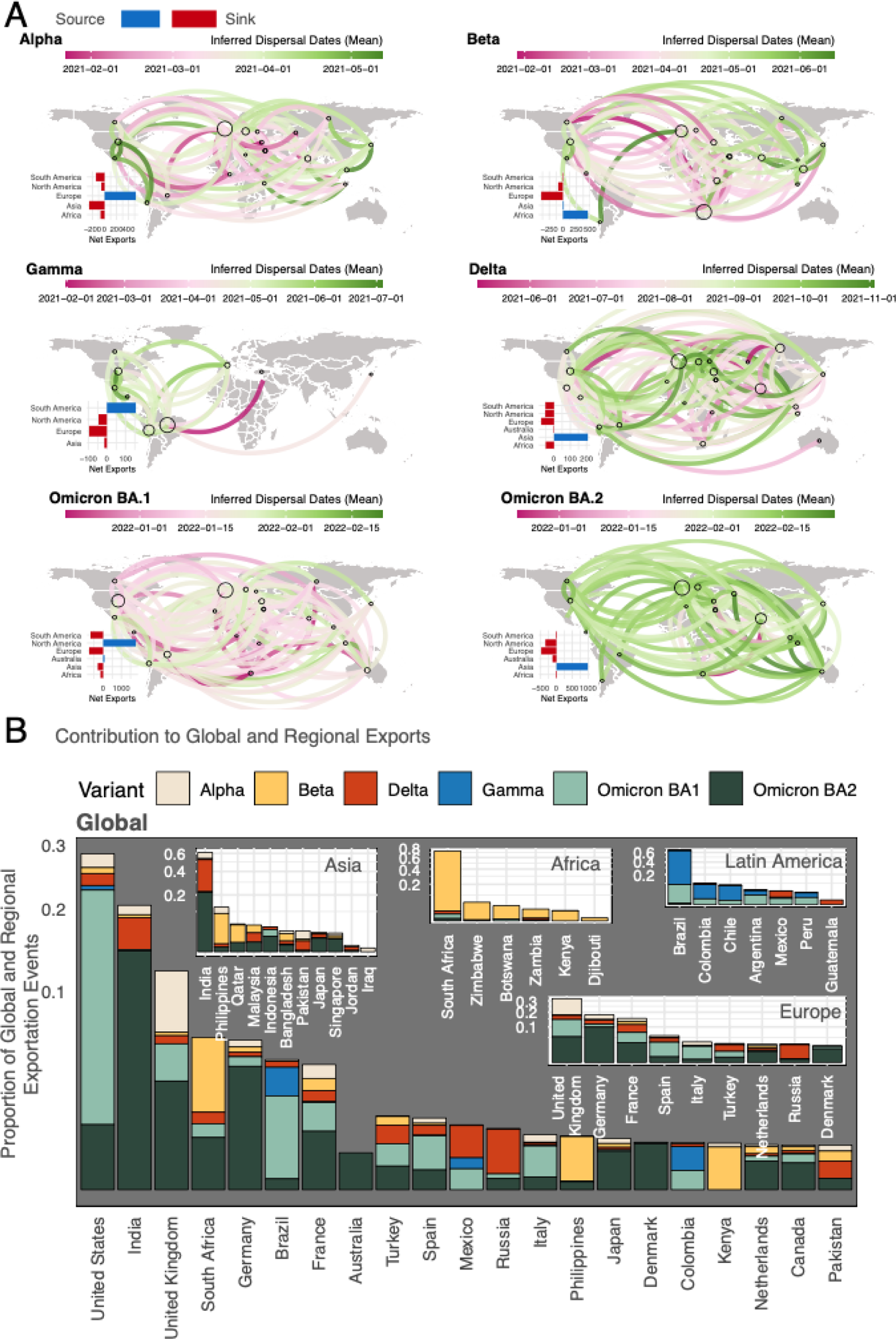
Spatiotemporal dispersal patterns of VOCs. A) Global dissemination and continental sourcesink dynamics for each VOC, determined from ancestral state reconstruction analysis. Virus lineage exchanges are aggregated at sub-continental level and curves linking any two locations are coloured according to the mean dates of all viral movements inferred along this route. The curves denote the direction of movement in an anti-clockwise direction. Circles are drawn proportional to the number of exports per location. Source and sink continents are determined by calculating the net difference between viral exportation and importation events. The absolute numbers of exportation and importation events for each continent per VOC are shown in Figure S5. B) Largest global and regional contributors to viral exports, stratified by VOC. Countries are shown here if they contribute >0.1% of exports globally or within Europe, or >0.5% within other regions.

Most Beta exportations were from southern Africa (∼1200 estimated exports; ∼48% of Beta exports) and around half of those were to locations within the same region (>600 exports) (Figure 1A). We also infer considerable Beta spread from southern Africa to Western Europe (∼300 exports) and then within Western Europe (∼400 exports). We infer that Asia was a net source of Beta along with Africa, with exports peaking after April 2021 (Figure S2), which is plausible given sizable Beta waves in some countries in Asia during that time (e.g. Bangladesh and Cambodia)^19^. We again observe a multi-stage process of Beta spread: during the earliest stages between late 2020 and early 2021, Beta was primarily exported from southern Africa into North America, Europe and Asia. Later, dispersal from Asia occurred mostly to Europe and North America, with minimal introductions to South America (Figure 1A). Gamma circulation was first detected in Brazil where it caused a large wave of infection between December 2020 and March 2021^3^. From there, Gamma was mainly exported to other South American countries (∼50% of Gamma exports). Only later, between May-June 2021, do we infer a few instances of Gamma spread from North America to Europe or from Europe back to the Americas (Figure 1A).

The global spread of Delta was marked by early exportations from the Indian subcontinent and Russia to other regions of Asia and all other parts of the world in the first half of 2021 (Figure 1A). Russia’s estimated contribution to international Delta exports (∼11% of global Delta exports in our study) is consistent with previous work inferring dispersal from there based on human mobility data^6^. In the second half of 2021, many more inferred dispersal events originated from Western Europe, including the UK (>1100 within Western Europe, >300 to Central Europe, and ∼30 to the USA, Brazil and the Middle East, respectively; Figure 1A, Figure S2, also in line with previous work^6^). While Western Europe demonstrated the largest absolute number of exportation events of the Delta variant, Europe still acted as an overall sink, and Asia remained a major net source when considering the net volumes of inbound as well as outbound Delta movements (Figure 1A).

We inferred the dispersal patterns for Omicron lineages BA.1 and BA.2 separately, given their genetic distinctiveness, until March 2022. Although both lineages were detected first in southern Africa around the same time, their dynamics of global dissemination differed. Consistent with the first major Omicron BA.1 waves occurring in southern Africa, we infer that the earliest exportation events of BA.1 originated from there during November and December 2021. Most Omicron BA.1 international lineage movements occurred within Western Europe (>2000) and we infer the timing of those transmissions to be centered around mid-January 2021 (Figure 1A). There was also considerable spread of BA.1 within the Americas (∼500 exports) and from North America to Europe (>800 exports) during the same time period. In comparison there were only 70 and 191 estimated exportation events from southern Africa to North America and Western Europe, respectively. We estimate that Omicron BA.2 early exportations from southern Africa were to the Indian subcontinent and to Europe, also starting in late November 2021. Germany, India and the UK were the three largest exporters of Omicron BA.2 overall, with 171, 170 and 100 inferred exports, respectively. Africa received approximately the same number of re-introductions of BA.1 and BA.2 as were originally exported from the continent (∼54 and ∼60 inferred exports versus ∼198 and ∼88 inferred re-introductions). We infer North America to be the major source of BA.1 and Asia of BA.2 (Figure 1A). Crucially, this means that the continent where these lineages were first reported did not act as a major global source of the VOC (Figure 1A, Figure S2). Although both lineages emerged around the same time in late 2021^8^, international dispersal of BA.2 occurred, on average, later than BA.1 (Figure 1A). This is expected given that, globally, BA.1 took off and fueled large epidemic waves before BA.2 did, and that BA.2 partially evades BA.1 immunity^20^ and therefore had a competitive advantage only after BA.1 had spread. Further, some countries were still experiencing large Delta waves when BA.1 and BA.2 were imported, potentially slowing their spread (e.g., Germany).

Exploring the mechanisms of variant expansion globally, we observe a strong correlation between the nationally recorded case incidence attributed to each variant and the inferred volumes of exports out of corresponding countries (Figure S3), although this is partly influenced by the case-sensitive genomic sampling strategy. We also observe a positive correlation between the connectivity of countries to global air travel networks and their inferred contribution to export numbers for all variants except for Beta and Gamma (Figure S3). This effect is likely a combination of lower international travel during the time of dispersal of Beta and Gamma, and their emergence in countries not on the backbone of international flight networks while Alpha caused a large outbreak in a well connected region. In fact, from December 2020 to March 2021, the total number of passengers out of the UK, South Africa and Brazil were ∼3.8million, ∼440,000 and ∼860,000 respectively, adding to the understanding of a larger geographical reach of Alpha compared to Beta and Gamma. As expected, this suggests that both case incidence and mobility play a role in determining the extent to which a country participates in global dissemination of variants, unless there is co-circulation of different variants in the world. In this case, local variant-specific incidence will be a better predictor of estimated variant exports from a certain country and mobility will only matter from regions where a variant is dominant (Figure S3).

### Quantifying regional and global dissemination hubs

Next, we investigated global and regional viral exports to identify hubs of international dissemination of different VOCs. Our results reveal that the United States (US) was the largest contributor to VOC exportations globally, responsible for close to 30% of all inferred VOC exports to other countries, followed by India, the UK, South Africa and Germany contributing roughly 20%, 12%, 6% and 5% of global VOC exports, respectively (Figure 1B). The share of contributions to international exportations is highly correlated with countries’ total air travel passenger volume (Spearman correlation □= 0.71, p < 0.001, Figure S4). However, we show that this role of important global hubs varied among VOCs. For instance, South Africa acted as a major global hub for viral exportation for the Beta variant. The US’s role was most visible for Omicron BA.1 (∼75% of its global exports), while India’s share of global exports was dominated by Delta and Omicron BA.2 (∼70% of its global exports), and the UK’s by Alpha (∼48% of its global exports). As with the dispersal patterns discussed above, the most important inferred contributors of Omicron dissemination globally were more proximal (from secondary locations) rather than distal (from southern Africa). These findings are well supported by reported epidemiological trends. For example, the US experienced large BA.1 waves in late November and December 2021 in metropolitan and highly connected cities on the East Coast (Washington D.C. and New York City). Similarly, a large number of BA.2 infections were reported from India after it had spread there from southern Africa^21^. We observe that countries that acted as major global hubs were also important in disseminating VOCs regionally (i.e. within the same continent). For example, Beta and Gamma were variants that mainly expanded regionally and we find that the dissemination of these variants in Africa and South America accounted for >50% of exportations in those regions (Figure 1B). A few countries also emerge as large regional hubs of viral exportations despite low global contributions: for example the Philippines and Pakistan in Asia, Colombia in South America, and Spain and Italy in Europe (Figure 1B), potentially linked to a combination of early seeding, large epidemics and passenger numbers.

The inferred networks of viral dispersal that we discuss here are dynamic, as shown by the heterogeneous patterns when comparing VOCs. These are likely influenced by localized epidemic sizes, human mobility, population immunity and viral variant phenotypes. However, an immediate implication of these findings is that the tendency of countries to act as important global or regional hubs of viral dissemination is dictated by local case incidence and the countries position in the hierarchical global air travel network^22^, rather than the location of emergence of variants (Figure S3).

### Role of First-Reporting Countries in Global VOC Dispersal

Following the emergence of each VOC, a variety of travel and passenger quarantine restrictions (Figure S6) have been put in place^23–25^. For example, travel restrictions were implemented against South Africa for 291 days after the discovery of Beta, against South America for 270 days following the discovery of Gamma, and then again for South Africa for around a month for Omicron^23^. Here, we investigate the inferred sources and timing of international introductions of VOCs from the place of first detection and contrast them to importations from all other locations (UK for Alpha, South Africa for Beta, Omicron BA.1 and BA.2, Brazil for Gamma and India for Delta).

The main finding for all VOCs is that even though VOC exports initially occurred mostly from the country of first reporting or presumed origin, this progressively shifted and more countries became sources of exports to other locations as incidence in other countries increased (Figure 2). We observe that this shift happened much more rapidly for Delta and Omicron compared to Alpha, Beta and Gamma (Figure 2). For Alpha, we find that by December 2020, roughly 100 days after the estimated date of emergence of this variant (TMRCA), around 20 countries had already received at least one inferred introduction of the variant (underestimate of real number of introductions) and were themselves acting as sources of exportations from established local transmission (Figure 2B). This suggests wide geographic dispersal and cryptic transmission by the time travel restrictions were implemented around December 2020 to control dissemination of Alpha. This pattern of gradual shift of the inferred source away from the presumed country of origin is similar for Beta and Gamma, the only peculiarity being the lower number of countries inferred to have received an introduction of Gamma. Of all introductions to other locations that we infer over time, the UK contributed 48% of Alpha, South Africa contributed 37% of Beta and Brazil 60% of Gamma (Figure 2A).

**Figure 2:**
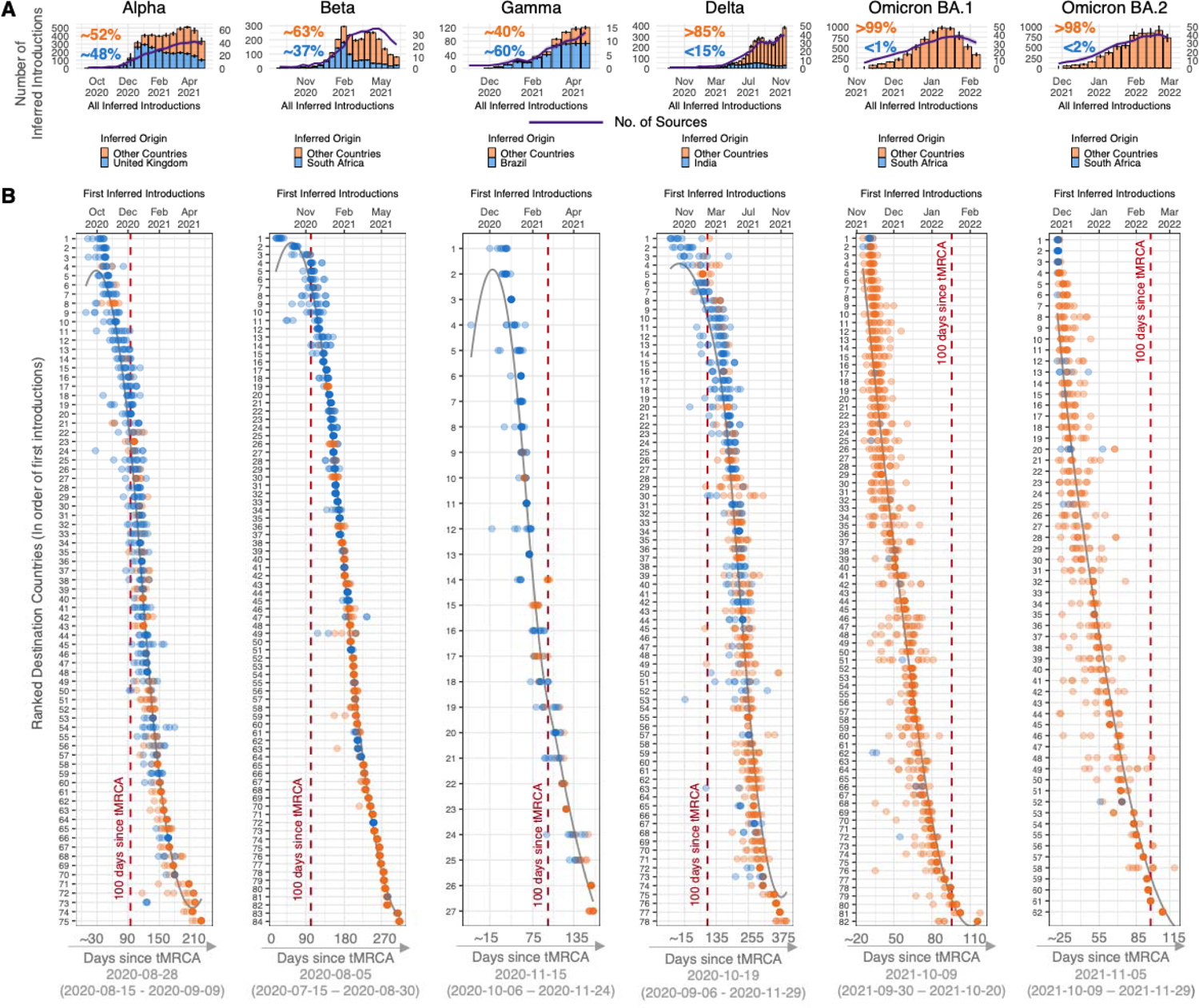
Inferred origins of global VOC dissemination events. A) Changes in proportions of all inferred introductions from the country of presumed origin for each VOC (bars), and the number of countries inferred to be acting as onward sources of each VOC (purple line, with scale in the second y-axis). Results shown are determined from 10 replicates of genome subsampling. B) Date of first inferred introduction per country coloured by location of origin, i.e. presumed origin (blue) or not (orange). The lines represent countries, which are ranked and labeled by the median of their dates of first introduction (from 10 replicates). The lower x-axis denotes the estimated median tMRCA and confidence interval range dates for each VOC, as reported in published studies^1–3,6,8,26^.

For Delta and Omicron BA.1 and BA.2 lineages, the pattern is different: India and South Africa, the presumed countries of origin of Delta and Omicron, respectively, very rapidly transitioned to being minor sources of both first and overall introductions of these VOCs to other countries. In the case of Delta, fewer than 15% of all introductions to other countries were attributed to India (Figure 2A). While India’s contribution to first and overall Delta introductions did not completely subside over time, it became heavily interspersed by the contribution of other countries as a source of Delta. The shift away from the presumed origin is even more marked for Omicron BA.1 and BA.2. Overall, we infer that South Africa was the source of fewer than 1% and 2% of BA.1 and BA.2 introductions globally (Figure 2A). We show that within the first week (early November) of BA.1 and BA.2 global dissemination, the first introductions to other countries were already originating from places other than South Africa. We also observe from the temporal reconstructions of these events that by the time travel restrictions were placed against southern Africa in December 2021, Omicron BA.1 exports could already be inferred from more than 30 other countries. Additionally, 100 days after estimated TMRCA of Omicron BA.1 and BA.2, we could already infer introductions into more than 80 and 60 countries, respectively, in stark contrast to the dispersal of the other VOCs during the same time frame (Figure 2B).

For all the variants investigated here, the results point to the diminishing importance through time to the international dispersal of VOCs from the first presumed origin. This shift was more rapid with Delta and even more so with Omicron, potentially due to increased transmissibility of these variants, as well as fewer restrictions on travel and fewer public health and social measures in many places, meaning higher and more sustained local transmission and thus more opportunity for onward spread. These conditions allowed these VOCs to reach other countries more rapidly, often even before first detection of the variants by sequencing and in all cases before any travel restrictions were implemented. The fact that this phenomenon is even more notable for Omicron could further be explained by a considerable increase in international travel volumes at the end of 2021 while travel bans against southern African countries were implemented very rapidly following the first report of Omicron emergence.

### Impact of international travel on VOC dispersal

The transmission of the SARS-CoV-2 virus was accompanied by major shifts in human mobility patterns throughout the pandemic^10,15,27^. In addition to national lockdowns restricting local movements and mixing, varying levels of air passenger travel restrictions were implemented in response to the initial emergence of SARS-CoV-2 and subsequent waves of transmission (Figure S6). In reconstructing the global dispersal of VOCs, the substantial decreases and more recent increases in international air travel might explain the variation in dispersal of VOCs alongside differences in immunity and vaccination. To examine how air travel has influenced the speed of dissemination of VOCs globally, we investigated global air travel passenger volumes between February 2020 to March 2022 and compared them to the speed of dispersal of VOCs in countries reporting VOCs using genomic data. This delay was quantified here as the number of days between the TMRCAs (median) of each VOC (Omicron BA.1 and BA.2 separately) from published studies (Supp Tab. S1) and the date of collection of the first sequenced VOC sample in each country (Source: GISAID).

We find that the Alpha, Beta and Gamma variants were first sampled for sequencing in countries around the world with a delay of 64 - 425, 95 - 300, and 48 - 251 days (5-95th percentiles), respectively, following their emergence (Figure 3A). On average, it took longer for countries to first sequence the Delta variant, with a delay of 45 - 336 days (5-95th percentiles) after its estimated date of emergence in October 2020^6^ (Figure 3A). The relatively longer delay between emergence and dates of sequencing can potentially be explained by the rapid spread of the Alpha, Beta and Gamma variants during that time in other countries and the relatively long period between emergence and rapid spread of Delta in India leading to global dissemination. In the case of Omicron, both the BA.1 and BA.2 lineages dispersed around the world much faster compared to the preceding VOCs, sampled in countries within just 7 - 98 and 28 - 186 days (5-95th percentiles), respectively (Figure 3A). This was likely strongly influenced by a three-fold increase in global air travel passenger volumes, to ∼60 million per month during the spread of Omicron, compared to ∼20 million per month a year before when the Alpha, Beta, Gamma and Delta variants disseminated (Figure 3A). Additionally, it is now well known that Omicron lineages were highly immune evasive, causing infections globally at much higher rates (see also next section).

**Figure 3:**
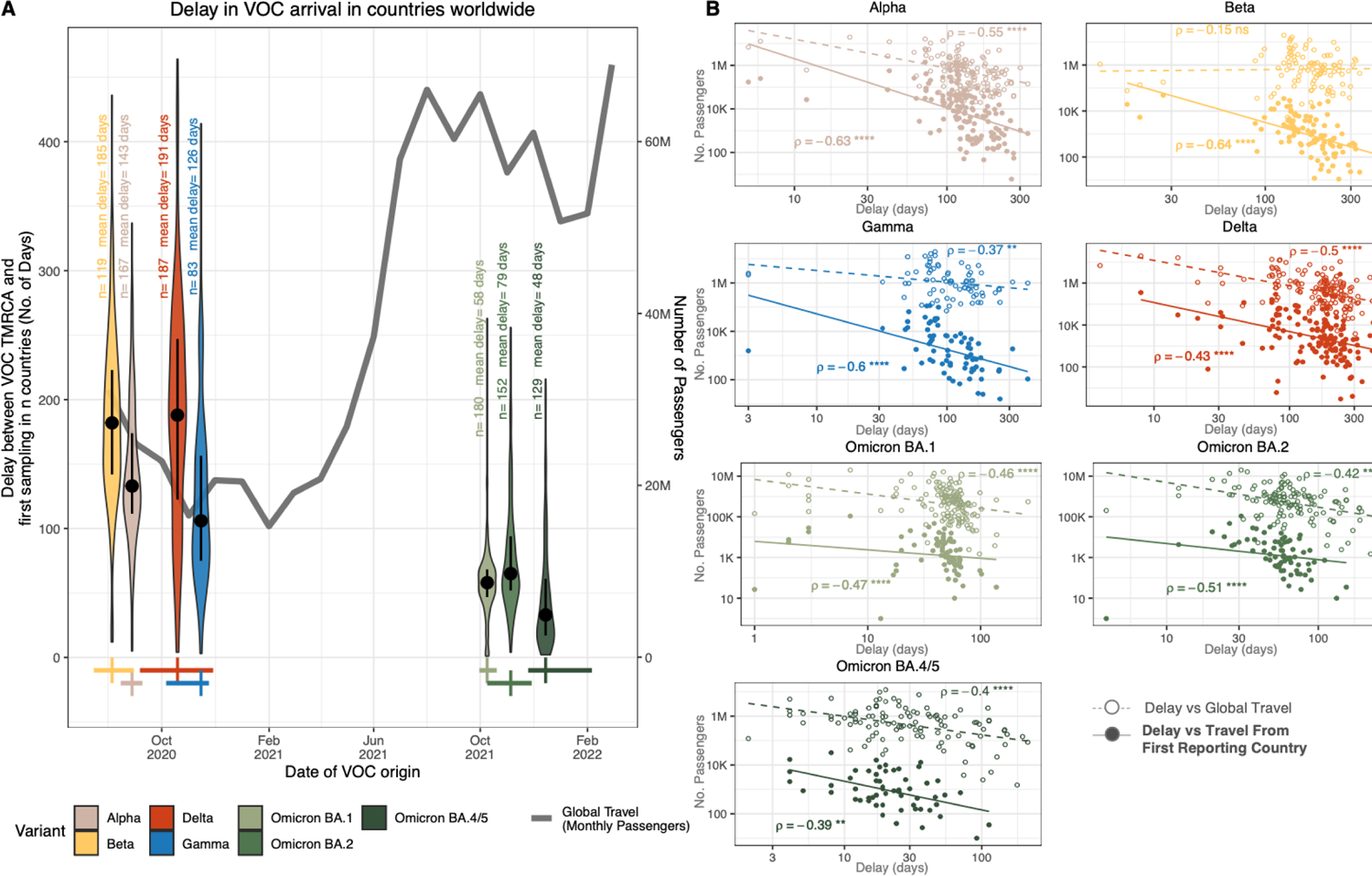
Impact of global air travel on VOC dissemination. A) Delay (number of days since TMRCA) of each VOC to be first sampled in countries around the world, and the total global monthly air passenger volumes from September 2020 to March 2022. The number of points and mean of each violin plot are indicated. The dot and error bars inside each group denote the median and interquartile range, respectively. Dates of VOC origin are taken as their published mean estimated dates of emergence (TMRCA), with crosses representing the median and high confidence range values^1–3,6,8,26^. The date of arrival of each VOC per country is taken to be the first sampling date of a sequenced case in GISAID (Date of access = 18 September 2022). B) Scatter plot and spearman rank correlations of either travel volumes from the first reporting country or total global travel with the delay in first sampling of VOCs in countries globally. Spearman rank correlation values are shown, with level of significance indicated.

To more specifically explore the relationship between global air travel and the delay in global VOC dispersal, we calculated pairwise correlations between sampling delays for each country for each VOC against the total incoming travel volumes to that country during the time of VOC dispersal, and against incoming travel volumes just from the presumed origin countries for each VOC. Overall, with the exception of Beta, we find negative and significant relationships between total travel volumes and delays to first sampling of VOCs in countries worldwide (Figure 3B), supporting the expected trend of higher travel volumes associated with lower VOC arrival delays, and vice versa (Figure 3B). We also find particularly strong negative correlations between these delays and travel volumes specifically from countries that first reported the respective VOCs, indicating that VOCs arrive in a country with less delay if that country has high travel volumes from the presumed country of origin of the VOC (Figure 3B). For Alpha, Beta and Gamma, total volume of travel from the UK, South Africa and Brazil, respectively, predicted better the delays in their first sampling in other countries compared to total global travel, pointing towards a lesser contribution of global transit hubs than for Delta and Omicron. For Delta and Omicron, the negative correlation with total global travel was as strong as travel from the presumed country of origin. This further supports previously discussed findings that locations other than the presumed origin quickly became exporters of viral lineages given high transmissibility of the variant. Delays in Gamma dissemination demonstrated slightly lower levels of correlation with total global travel (Figure 3B). Given the largely regional reach of Gamma compared to others analyzed here, this finding can be explained by the greater importance of air travel connection specifically to Brazil, the presumed country of origin. Further modeling work is needed to quantify the relative contributions of the location of origin to the epidemic dynamics elsewhere as compared to the global air travel network^22^. We also reperformed the analysis using dates of first inferred introduction of variants globally, rather than dates of first sequence, and our results are comparable (Figure S7).

### Global Epidemic and Variant Dynamics

In addition to distinct dispersal dynamics described in previous sections, VOCs emerged in heterogeneous epidemiological landscapes globally (Figure 4) where differences in the epidemic intensity around the world were further exacerbated by uneven diagnostic testing rates (Figure 4A, Figure S8), distinct levels of population immunity as the pandemic progressed (either vaccine- or infection-acquired), and variation in geographical and temporal drivers of transmission. Despite underreporting of infection numbers^28^, combining death, genomic surveillance and testing data provides qualitative insights into the differences in epidemic waves across continents (Figure 4). While Alpha, Beta and Gamma expanded regionally, Delta and Omicron swept across the globe becoming dominant worldwide in middle-late 2021 and early 2022, respectively (Figure 4A).

**Figure 4:**
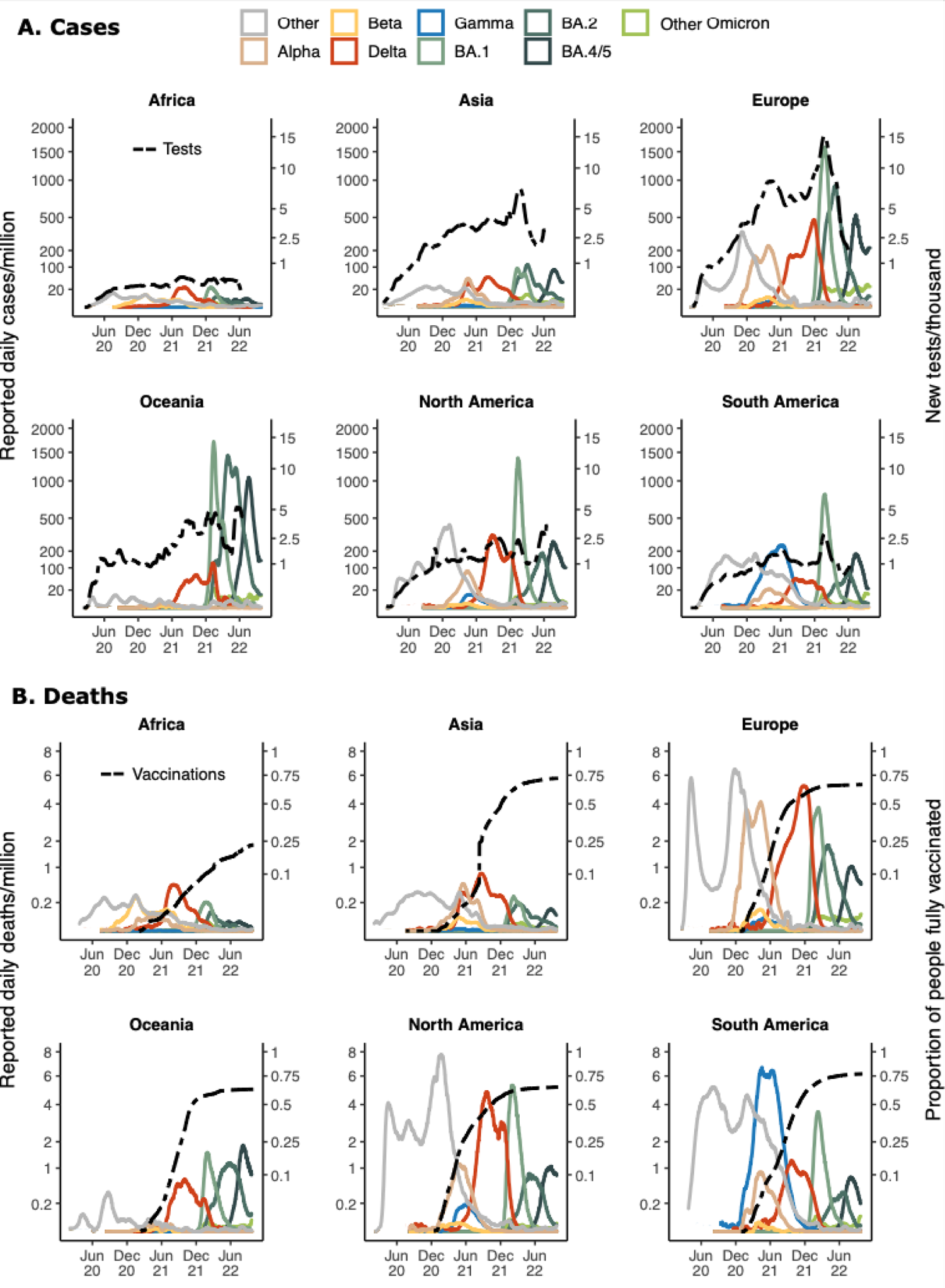
Continental epidemiology of SARS-CoV-2 cases, mortality, testing and vaccination. A) Th progression of daily reported cases per continent from February 2020 to October 2022 (log scale, first y-axis). The 7-day rolling average of daily reported case numbers is coloured by the inferred proportion of variants responsible for the infections, as calculated by genomic surveillance data (GISAID date of access: 1 October 2022) averaged over 20 days. The line shows the 7-day rolling average of the number of daily tests per thousand population per region (scale shown in the second y-axis) aggregated for countries for which this data is available for each continent. B) The 7-day rolling average of daily reported deaths coloured by the inferred proportion of variants, as calculated for case data, with an assumption of time lag of 20 days between infection and death applied (See more details in Materials and Methods). The dashed line displays the proportion of people fully vaccinated per region (scale on second y-axis) where those that received all doses prescribed by the initial vaccination protocol are considered fully vaccinated.

Throughout the different waves of infection, we observe a marked difference in reported mortality in Africa, Asia and Oceania compared to Europe, South and North America (Figure 4B). This is likely due to a combination of underdetected death counts in Africa and Asia, as suggested by high levels of modeled excess deaths^28^, and low virus circulation in Oceania due to prolonged border closures in earlier stages of the pandemic^29,30^. Despite high vaccination coverage and accelerated booster rollout during the emergence of BA.1 (Figure 4B), case incidence increased rapidly (Figure 4A) in the context of relaxing non-pharmaceutical interventions (NPIs) across the world. The high number of cases meant considerable mortality especially across a few locations with no immunity, due to a combination of low population exposure to the virus and low vaccination rates in high risk groups, for example in Hong Kong^31^. In absolute numbers, Oceania, North and South America all experienced higher mortality due to Omicron BA.1 than Delta. Vaccine inequity exacerbated the inability to protect even high risk groups in many places at this stage, particularly considering low vaccination coverage in Africa (Figure 4B).

## Discussion, limitations and future direction

Our study shows that SARS-CoV-2 VOCs (Alpha, Beta, Gamma, Delta, and Omicron BA.1 and BA.2) disseminated around the world according to different spatial source-sink dynamics and that generally, global travel hubs were important contributors of viral exportations. The international spread of Delta and Omicron were substantially different to the dissemination of Alpha, Beta and Gamma. The dispersal of Delta and Omicron was generally more multi-focal, with multiple regions contributing to its global invasion. Also, Australia contributed to viral exchanges for Delta and Omicron (unlike Alpha, Beta and Gamma) consistent with published reports of Delta introductions despite quarantine measures, and the opening of borders for non-Australian citizens prior to the Omicron wave^32–34^. These differences reflect both the distinct global landscapes at two stages of the pandemic and the intrinsic variant characteristics. Alpha, Beta and Gamma perhaps circulated in restricted regional locations, whereas Delta and Omicron dominated infections in a global sweep. The period during which Alpha, Beta and Gamma emerged was characterised by lower global mobility and widespread travel restrictions, whereas the gradual lifting of such restrictions facilitated the more rapid and widespread Delta and Omicron dissemination^6^.

We also investigated the role of the presumed origin location of each VOC (the countries that first reported each variant) in the global dispersal of these viruses. While we infer that the UK, South Africa and Brazil were the source of the majority of global exportations of Alpha, Beta and Gamma (all >35%), we find that for Delta and Omicron, the contribution of the presumed origin was much smaller (<15%). We observe differences in the speed at which countries that are not the presumed origin became exporters of the VOC. For Delta and Omicron, this rapid transition could be attributed to a mix of increased transmissibility and increased global air travel during the times when they spread. Our results should be viewed in the context of countries’ epidemiological landscape. The pattern that we present, however, highlights that locations with high case incidence and global connectivity have the potential to become major contributors of variant exportations if early seeding of viral variant outbreaks are not controlled.

SARS-CoV-2 variants continue to emerge and spread worldwide, as seen most recently in the case of the Omicron BA.4 and BA.5 lineages^26^. In this context, our findings have some general implications for public health. First, we show that once a variant has been established in multiple countries, continued international spread may almost be inevitable. When specific routes are shut through travel restrictions, other locations become responsible for a greater share of global dissemination. This indicates that in the case of emerging variants, especially those with enhanced virulence and waning immunity, actions to control or mediate the effects of virus transmission should be undertaken everywhere. Second, our results indicate that travel restrictions, especially targeted ones, are often implemented after initial imports have already come into other countries, especially for more transmissible variants, as discussed by others before^25,35^. To limit the extent of local transmission, a combination of measures are necessary to contain spread, including testing at arrival, antigen-testing before large gatherings, isolation while infectious, and vaccination, among others^36^. Lastly, as global air travel and human mobility rebound towards prepandemic levels and beyond, new variants are likely to reach secondary countries much faster, potentially before being identified by genomic surveillance. Despite the massive effort of genomic sequencing globally, the nature of respiratory viruses like SARS-CoV-2, and especially highly transmissible VOCs, means that testing and genomic surveillance will often struggle to detect a new variant before there is ample opportunity for wide dispersal^37^. This makes targeted travel restrictions increasingly redundant, and continued investment and innovation in robust, fast, and systematic diagnostic and surveillance programs is crucial for current and future pathogens. For example, targeted genomic surveillance can be informed by a location’s position in the hierarchical global air travel network.

The findings presented here are derived primarily from a phylogeographic analysis and have several limitations. Our genomic sampling was informed by the timing and size of epidemics per country to avoid over-representing countries with more sequencing, and to focus inferences on a subset of sequences that most accurately represents routine surveillance of local transmission, rather than possible targeted sequencing of travel-related cases or over-sequencing at the beginning of waves (Materials & Methods). However, we cannot rule out remaining biases in our datasets, particularly those associated with uneven testing rates and case reporting globally. With this method of sampling, it is also not possible to unambiguously identify the very first introductions of VOCs, especially if associated with cryptic transmission for a prolonged time before a marked increase in variant related cases. We also rely on national genomic surveillance data to scale reported cases to variants and we recognize biases linked to that, including possible uneven representation of incidence from subregions of countries, and non-uniform sequencing proportions at various stages of the pandemic. Furthermore, the global and continental view taken in our analysis will obscure fine scale heterogeneity within countries where epidemiological trends can differ.

In order to best represent VOC epidemics worldwide, we performed phylogenetic inferences on datasets of roughly 20,000 genomes for each VOC, in replicates of ten. Given the size of these datasets we were not able to employ full Bayesian phylogeographic reconstruction methods given availability of computational resources. This would mostly affect the precise timing of inferred ancestral state changes per country, however the very dense sampling through time puts some strong temporal constraints on when a state change can occur. Finally, our analysis focuses on the spatiotemporal invasion aspect of the global reach of VOCs, and does not attempt to study the impact on epidemic growth in countries that do receive an introduction of these VOCs. Therefore, we cannot make a causal claim between the size of the epidemic wave in the destination country and the number of viral introductions and travel volumes. Other studies have shown that local measures in the destination country influence the control of waves more than the number of seeding events^6^. Future work to document the extent of impact of the global spread of VOCs could additionally consider local contexts in relation to VOC characteristics, including population immunity profile, whether acquired through vaccination or infection waves of particular variants, parameters of waning immunity related to the duration of time from the last wave, and local control measures.

By systematically analyzing large representative datasets for each variant, this study highlights the role of global and regional hubs to viral dispersal while contrasting it to targeted travel restrictions towards the presumed origins of VOCs. Using travel data, we discuss that novel emerging VOCs with a clear transmission advantage are likely to spread much faster around the world given today’s increase in travel volumes. Overall, the global-scale spatiotemporal invasion patterns described here provide an opportunity to integrate knowledge of viral exportation and importation dynamics into our collective understanding of pandemic progression. This will be critical both in managing the upcoming stages of global SARS-CoV-2 transmission and within preparedness plans for future epidemics.

## Data Availability

The findings of this study are based on sequences and metadata associated with a total of XXX sequences available on GISAID up to November 19, 2022, via gisaid.org/EPI_SET_XXX. Custom data sources and scripts to reproduce the results of this study are publicly shared on GitHub (https://github.com/CERI-KRISP/SARS_CoV_2_VOC_dissemination). The repository contains all of the time scaled ML tree topologies, annotated tree topologies as well as custom data analysis and visualization scripts. Other datasets and pipelines used in this study are openly available and described in the Materials and Methods section.

## Acknowledgments

We gratefully acknowledge all data contributors, i.e., the Authors and their Originating laboratories responsible for obtaining the specimens, and their Submitting laboratories for generating the genetic sequence and metadata and sharing via the GISAID Initiative, on which this research is based. In Addition, we gratefully acknowledge all sources of funding associated with this work. In particular, KRISP and CERI is supported in part by grants from the Rockefeller Foundation (HTH 017), Abbott Pandemic Defense Coalition (APDC), the African Society for Laboratory Medicine, the National Institute of Health USA (U01 AI151698) for the United World Antivirus Research Network (UWARN) and the INFORM Africa project through IHVN (U54 TW012041), H3BioNet Africa (Grant # 2020 HTH 062), the South African Department of Science and Innovation (SA DSI) and the South African Medical Research Council (SAMRC) under the BRICS JAF #2020/049 and the World Bank (TF0B8412). M.U.G.K. acknowledges support from a Branco Weiss Fellowship, Reuben College Oxford, Google.org, the Foreign, Commonwealth and Development Office and Wellcome (225288/Z/22/Z), The Rockefeller Foundation, and from the European Union Horizon 2020 project MOOD (grant agreement number 874850). O.G.P. and M.U.G.K. acknowledge support from the Oxford Martin School. N.R.F. acknowledges support from Wellcome Trust and Royal Society Sir Henry Dale Fellowship (204311/Z/16/Z), Bill and Melinda Gates Foundation (INV-034540) and Medical Research Council-Sao Paulo Research Foundation (FAPESP) CADDE partnership award (MR/S0195/1 and FAPESP 18/14389-0).

The content and findings reported herein are the sole deduction, view and responsibility of the researcher/s and do not reflect the official position and sentiments of the funding agencies.

## Authorship contribution

Conceptualization: H.T, E.W, M.U.G.K, T.d.O

Methodology & Data Analysis: H.T, E.W, A.F.B, D.M, M.M, M.G, K.K, C.H, I.B, J.E.S, J.L.H.T, J.P, J.S.X, D.d.S.C, F.R

Supervision: N.R.F, M.U.G.K, T.d.O

Writing – original draft: H.T, E.W, R.J.L, D.M, M.U.G.K

Writing – review & editing: H.T, E.W, M.U.G.K, C.B, O.G.P, T.d.O, R.J.L, N.R.F,

## Methods

### Epidemiological Data and Genomic Prevalence

We analyzed trends in daily numbers of cases of SARS-CoV-2 and reported deaths for each continent up to 1 October 2022 from publicly released data provided by the Our World in Data repository (https://github.com/owid/covid-19-data/tree/master/public/data). To provide a comparable view of epidemiological dynamics over time in different continents, the variable under primary consideration for Figure 4 was ‘new cases per million’ and ‘new deaths per million’. Genomic metadata was downloaded for all entries on GISAID for the same time period (date of access: 1 October 2022). From this, information extracted from all entries for this study included: date of sampling, continent of sampling, viral lineage and clade.

We calculate the rolling average of daily case and deaths numbers for each variant by inferring the daily proportion of variants responsible for infections, as calculated by genomic surveillance data on GISAID. A lag time of 17.4 to 24.7 days (median = 20) was applied between the calculated genomic prevalence and recorded deaths, as reported in published literature^38^. A smoothing factor of 20 was used to calculate the rolling averages for case numbers, but the smoothing factor used of death numbers varied according to the consistency of the data, as follows: Africa k = 20, Asia k = 20, Europe k = 14, Oceania k = 22, North America k = 14, South America k = 22.

Vaccination statistics per continent and testing and positivity datasets per country were also obtained from Our World in Data. Overall continental data is not available in the dataset for the testing and positivity rate, and was therefore calculated from countries for each continent with available data. The proportion of people fully vaccinated per region was calculated with the number of people fully vaccinated, defined as having received all doses prescribed by the initial vaccination protocol, and the population size data provided by Our World in Data.

### Genomic sampling

Due to the sensitivity of ancestral state reconstruction methods to sampling bias, we performed our selection of genomic sequences in a careful manner to minimize biases to an extent that sampled datasets broadly reflect global reported case counts. From the complete set of entries for each VOC available on GISAID (Date of access: 17 September 2022), our subsampling strategy selected sequences to correspond to timeframes of global circulation of the respective VOCs, and in proportion to recorded cases in different countries. We used a previously described method *subsampler* (https://github.com/andersonbrito/subsampler)^39^ to produce such globally representative subsets for each VOC. This method subsamples sequences per country based on case counts over the study period to ensure the random sample is both geographically, temporally and epidemiologically representative. In short, *subsampler* requires the sequence metadata for the complete dataset from which the subsampling occurs, along with a case count matrix, which we scaled for each VOC to their estimated prevalences for different countries based on GISAID data. This subsampling scheme was performed ten times using ten unique random number seeds for each VOC to produce ten randomly sampled genomic datasets per VOC. Subsampling within this scheme is performed using a baseline function, which represents the proportion of cases that the user wishes to sample. This was changed accordingly to produce datasets of approximately 20,000 sequences for each VOC. Due to the genetic distance between the BA.1 and BA.2 Omicron variants, these two sub-lineages were split in downstream analyses and in this subsampling procedure. To minimize the potentially confounding effects of recombination on downstream phylogenetics-based analyses (which assume sequences are evolving in the absence of recombination), potential recombinant sequences were detected in the BA.1 and BA.2 subsets using RDP5.23^40^. Specifically each of the ten BA.1 and BA.2 specific datasets were analyzed using default RDP options other than that sequences were considered to be linear. Sequences flagged for signs of recombination with an associated P-value of 0.05 or lower were removed from the datasets. The final datasets for each VOC replicate contained the following numbers of sequences, corresponding to the specified date ranges to match relevant periods of circulation of each variant:

Dataset date ranges (sampling dates):

Alpha: n = 21,280, dates = 2020-10-19 - 2021-07-11

Beta: n = 22,669, dates = 2020-08-19 - 2021-08-07

Gamma: n = 21,331, dates = 2020-09-11 - 2021-09-22

Delta: n = 17,463, dates = 2020-09-07 - 2021-12-23

Omicron BA.1: n = 18,732, dates = 2021-11-17 - 2022-03-08

Omicron BA.2: n = 18,766, dates = 2021-11-27 - 2022-03-15

### Phylogenetic reconstruction

For each of the ten replicates per VOC, we produced time scaled tree topologies and performed discrete ancestral state reconstruction (of locations) to infer the global dissemination of each variant. Sequences were aligned using NextAlign^41^ and Maximum-likelihood tree topologies were inferred using FastTree v.2^42^ under a GTR model of nucleotide substitution. The resulting tree topologies were inspected for temporal molecular clock signals using the *clock* functionality of TreeTime^43^. ML-trees were then transformed into time scaled phylogenies in TreeTime using a standard mutation rate of 0.0008 substitutions per site per year and a standard clock deviation of 0.0004, and using the --confidence flag to get the 90% maximum posterior lower and upper bounds of divergence time estimates and confidence in state transitions in downstream *mugration* analysis. (Using a standard mutation rate resulted in inferences with better confidence compared to using an adjusted mutation rate for each VOC as determined in a root-to-tip regression analysis (Figure S9)). Outlier sequences that deviated from the strict molecular clock assumption as flagged by TreeTime were removed with the Ape package in R^44^ until a good time scaled phylogeny was obtained. The *mugration* package extension of TreeTime was then used to map discrete country locations to tips and infer country locations for internal nodes under a GTR model. Finally, a custom python script (available in our GitHub repository: <https://github.com/CERI-KRISP/SARS_CoV_2_VOC_dissemination>) was used to count the number of state changes over the span of the tree. State transitions that occurred prior to the earliest known tMRCA for each VOC and sub-variant were discarded to minimize the counting of transitions belonging to deep nodes with low confidence

### Source sink dynamics and viral movement patterns quantification

From the above ancestral state reconstruction data, phylogeographic maps and source sink dynamics were calculated and plotted using custom R scripts (available in our GitHub repository), as follows. Each recorded location state change is time stamped in decimal dates and annotated with origin and destination countries. Each of those state changes are further annotated with their corresponding continental and sub-continental groupings. Volumes of viral exports and introductions are calculated by aggregating the replicates for each dataset and the mean is considered either per country, continent, sub-continental region and specific to each VOC. Each state change is also annotated to be either a global (between two locations on different continents) or regional viral exchange (between two locations on different continents). Source and sink dynamics are estimated by calculating the net difference between the numbers of exports and introductions for a specific location; a location is determined to be a net source if the number of exports of a variant exceeds the number of introductions. The phylogeographic maps are constructed by linking sub-continental regions with curved lines going anti-clockwise in the direction of the curve. Each curved line is coloured by the mean date of state changes occurring along that specific link.

The speed of VOC arrival in different countries is calculated as the delay (number of days) between the estimated dates of emergence of each VOC from published literature (Supp Table S1) and either the first sampling dates of genomes per VOC and per country as reported on GISAID (Date of access: 18 September 2022) or the first inferred introduction from our ancestral state reconstruction data. Pearson correlations are calculated between this delay and mobility into different countries by considering either the total volume of passengers into each country, or the volume of passengers from either the UK (for Alpha), South Africa (for Beta and Omicron), Brazil (for Gamma), and India (for Delta) into each country.

All data visualization was generated through the ggplot package in R^45^.

### Air Travel Data

We evaluated travel data generated from the International Air Transport Association (IATA) to quantify passenger volumes originating from international airports during the specified time periods (reported below). IATA data accounts for ∼90% of passenger travel itineraries on commercial flights, excluding transportation via unscheduled charter flights (the remainder is modeled using market intelligence). Correlations with air travel passenger volumes were calculated using the Spearman rank correlation method, and reporting levels of significance.

Relevant travel periods:

Alpha: September 2020 - March 2021, Origin: UK

Beta: September 2020 - March 2021, Origin: South Africa

Gamma: November 2020 - May 2021, Origin: Brazil

Delta: September 2020 - September 2021, Origin: India

Omicron BA.1: November 2021 - March 2022, Origin: South Africa

Omicron BA.2: November 2021 - March 2022, Origin: South Africa

Omicon BA.4/BA.5: November 2021 - March 2022, Origin: South Africa

Global travel dataset: February 2020 - March 2022

## Supplemental Figures

**Figure S1.**
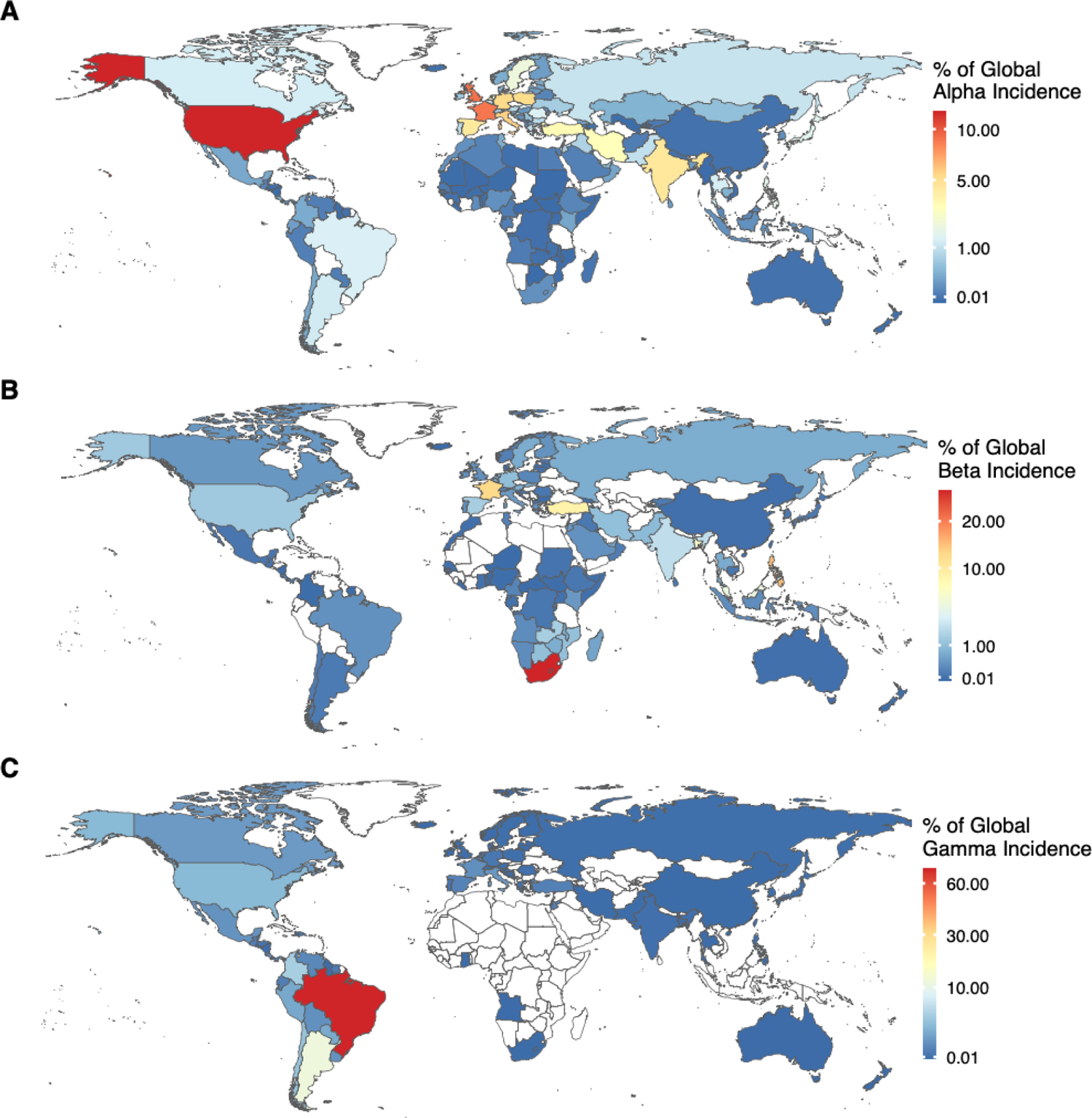
Alpha, Beta Gamma global distribution. Maps show countries coloured by their share of total global Alpha (A), Beta (B) or Gamma (C) incidence.

**Figure S2.**
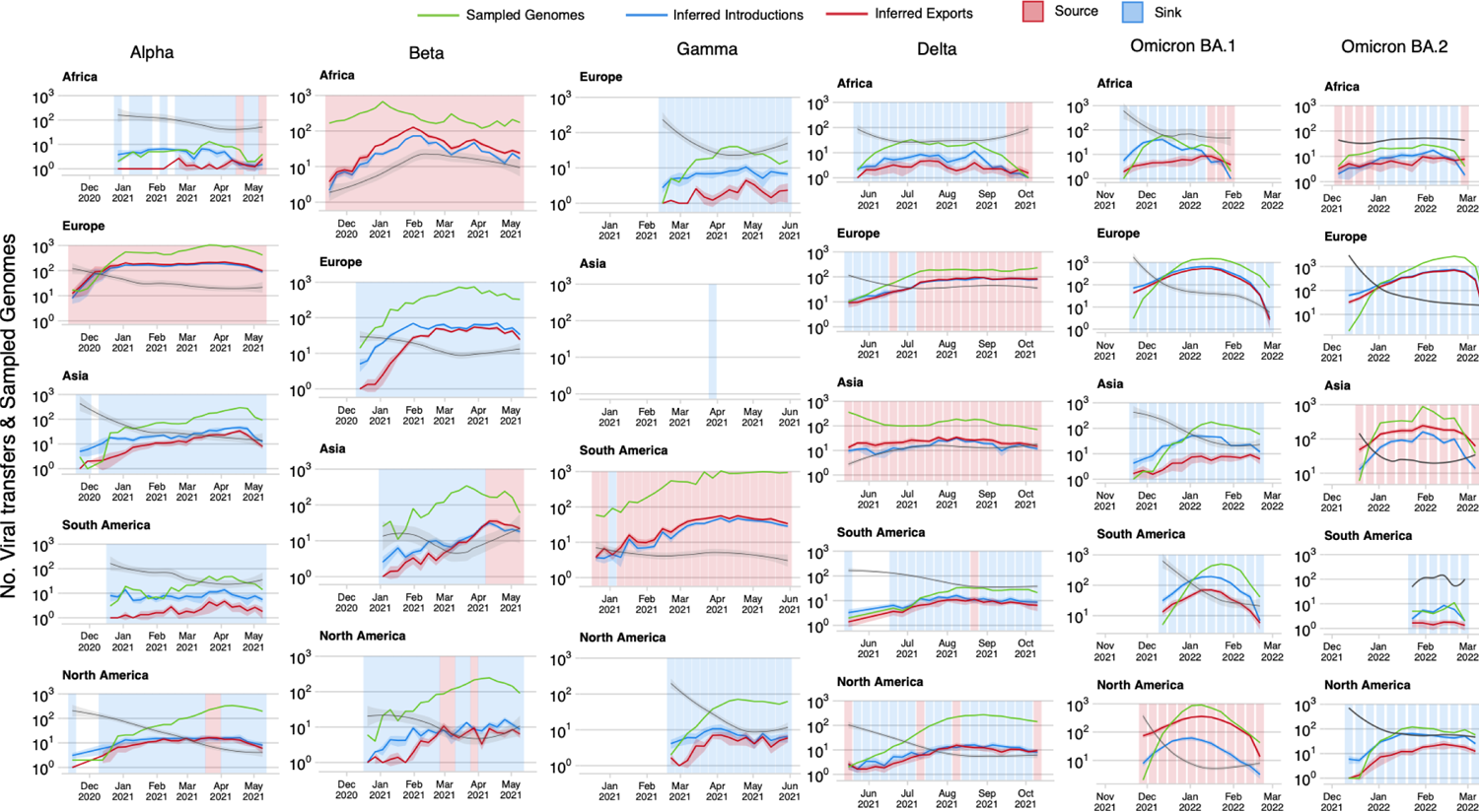
Viral sampling, introductions and exportations for various VOCs over time. Graphs show the time-varying progression of the numbers of sampled genomes in our analysis compared to the numbers of inferred introductions and exportations for Alpha, Beta, Gamma, Delta, Omicron BA.1 and Omicron BA.2 per continent.

**Figure S3.**
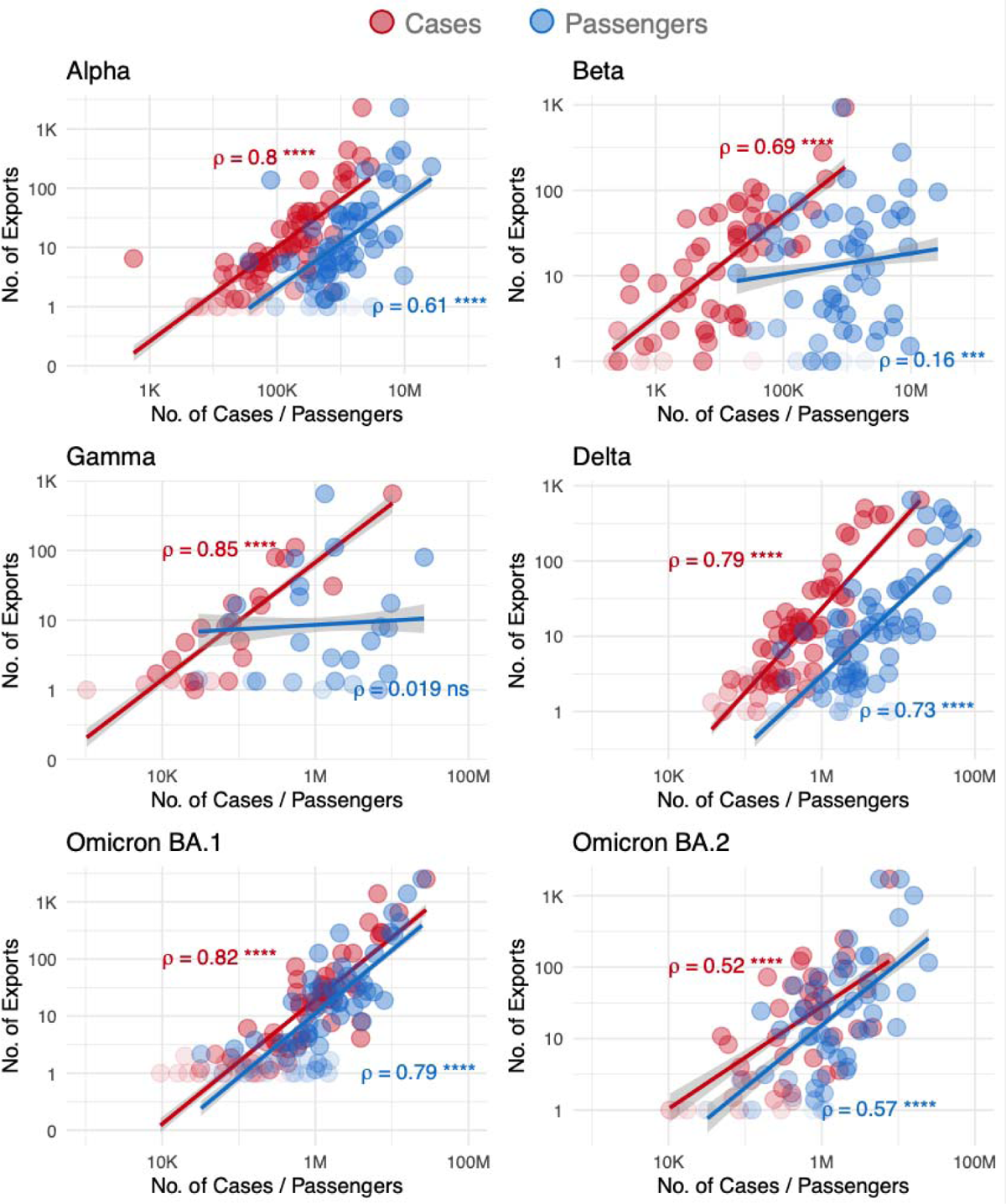
Correlations of Incidence and Travel to inferred VOC exportation numbers. Graphs show scatter plots and regression lines denoting the numbers of variant-specific cases, volumes of air travel passengers and inferred numbers of VOC exportations for each country. Spearman rank correlation values are shown, with level of significance indicated.

**Figure S4:**
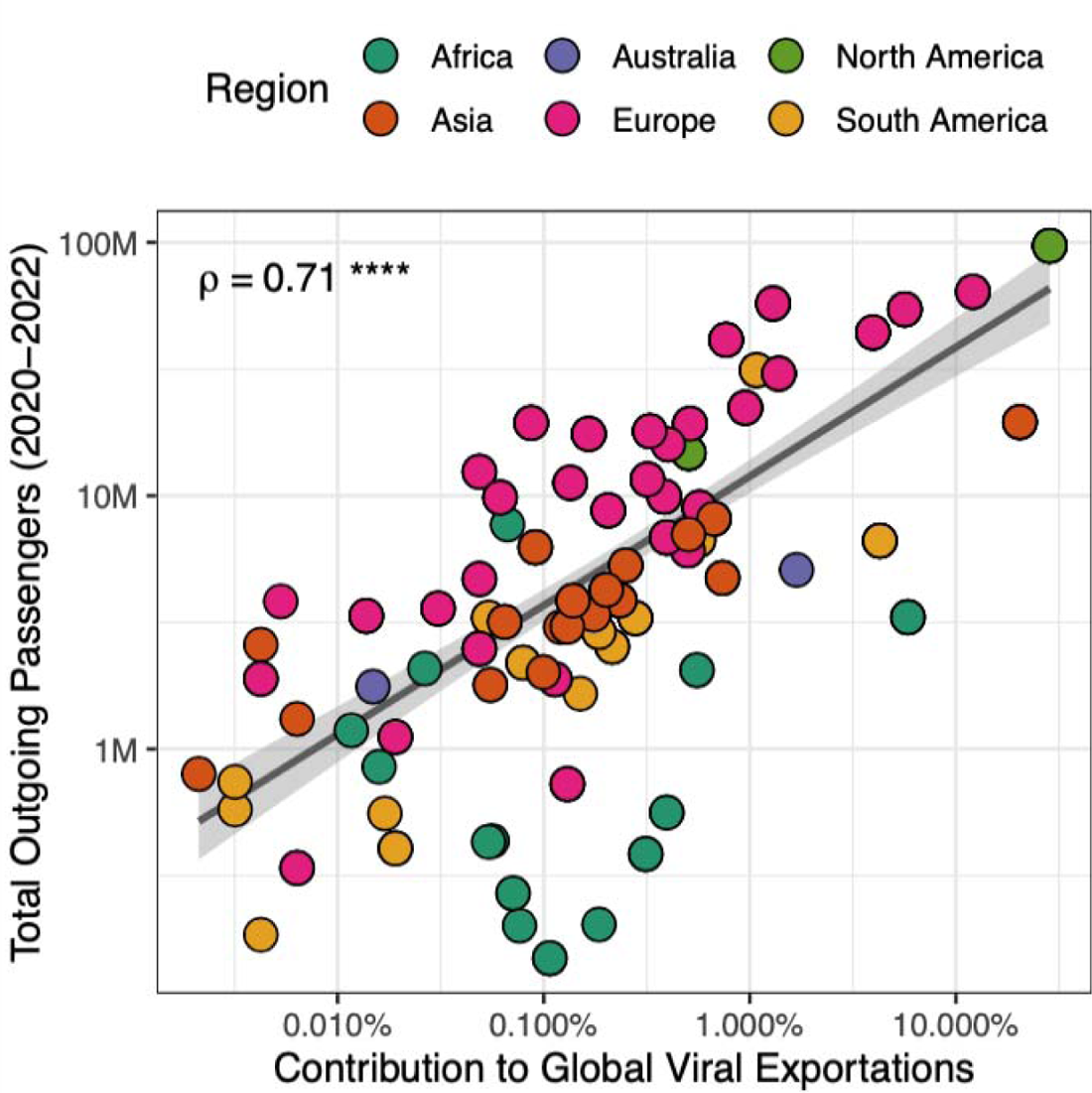
Correlation of Contribution of Global Viral Exportation Events and Outgoing Travel from Countries. Graph shows a scatter plot and regression line denoting the share of each country’s contribution to global numbers of inferred exportations for all VOCs and the total number of outgoing air travel passengers from 2020-2022. The spearman rank correlation value is shown, with level of significance indicated.

**Figure S5:**
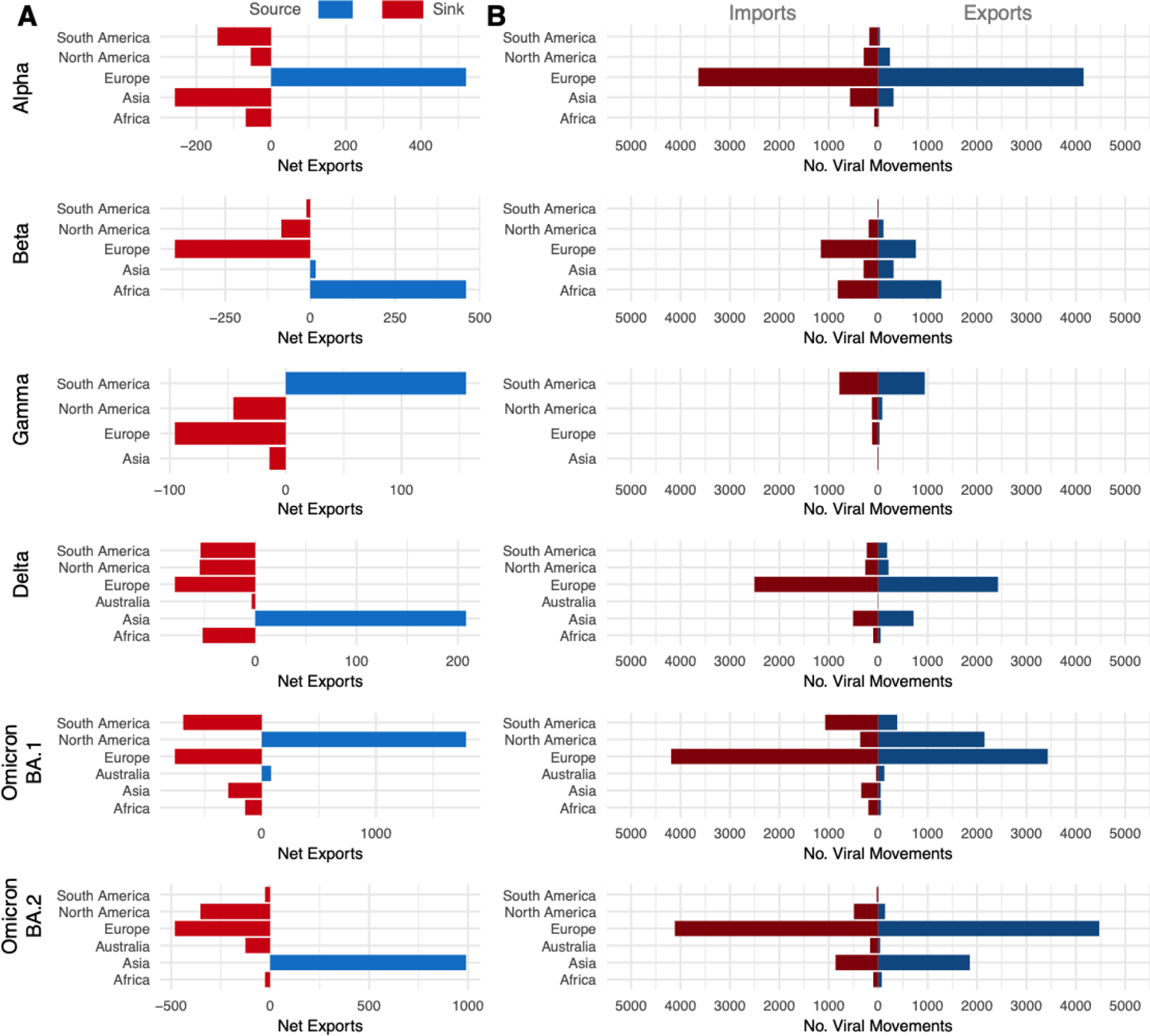
VOC imports and exports numbers per continent. A) The net difference between viral exportation and importation events. B) The absolute numbers of exportation and importation events for each continent per VOC.

**Figure S6:**
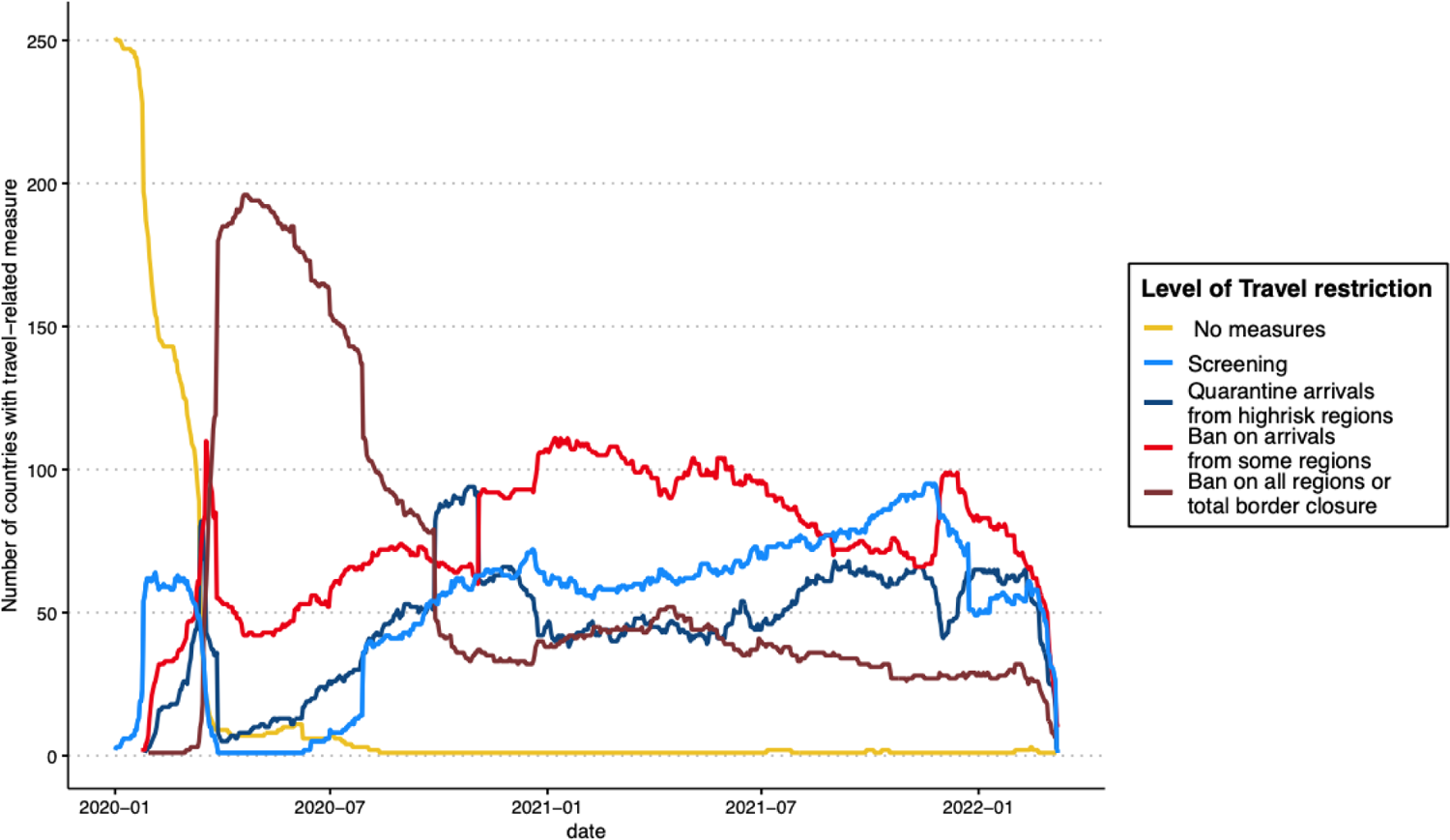
Air travel restrictions. The graph shows the time-varying numbers of countries implementing various levels of travel-related restrictions from January 2020 to March 2022.

**Figure S7:**
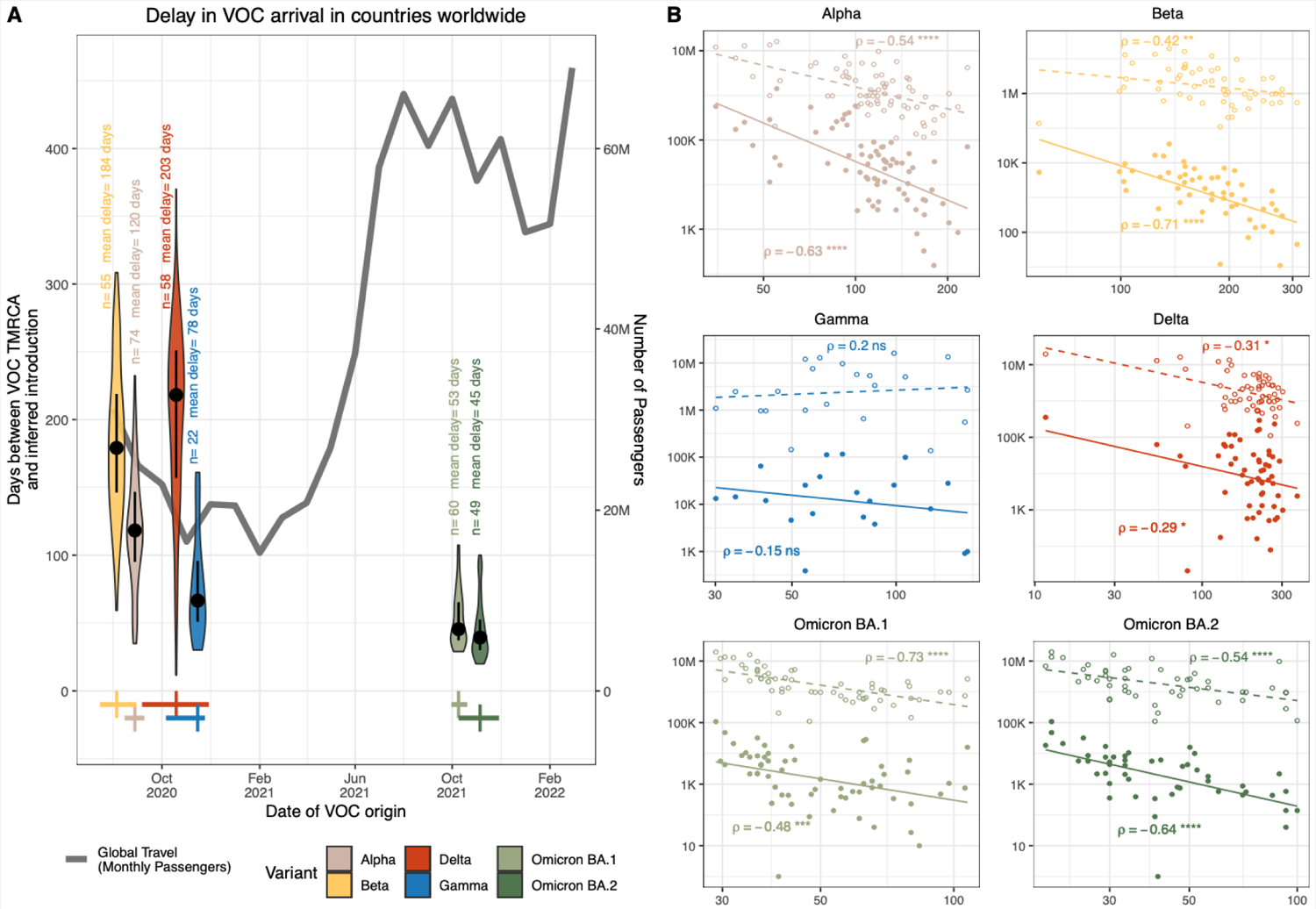
Impact of global air travel on VOC dissemination. A) Delay (number of days since TMRCA) of each VOC to first reach other countries around the world (arrival dates inferred as date of first introduction in phylogeographic analysis), and the total global monthly air passenger volumes from September 2020 to March 2022. The number of points and mean of each violin plot are indicated. The dot and error bars inside each group denote the median and interquartile range, respectively. Dates of VOC origin are taken as their published mean estimated dates of emergence (TMRCA), with crosses representing the median and high confidence range values^1–3,6,8,26^. B) Scatter plot and spearman rank correlations of either travel volumes from the first reporting country or total global travel with the delay in first introduction of VOCs in countries globally. Spearman rank correlation values are shown, with level of significance indicated.

**Figure S8.**
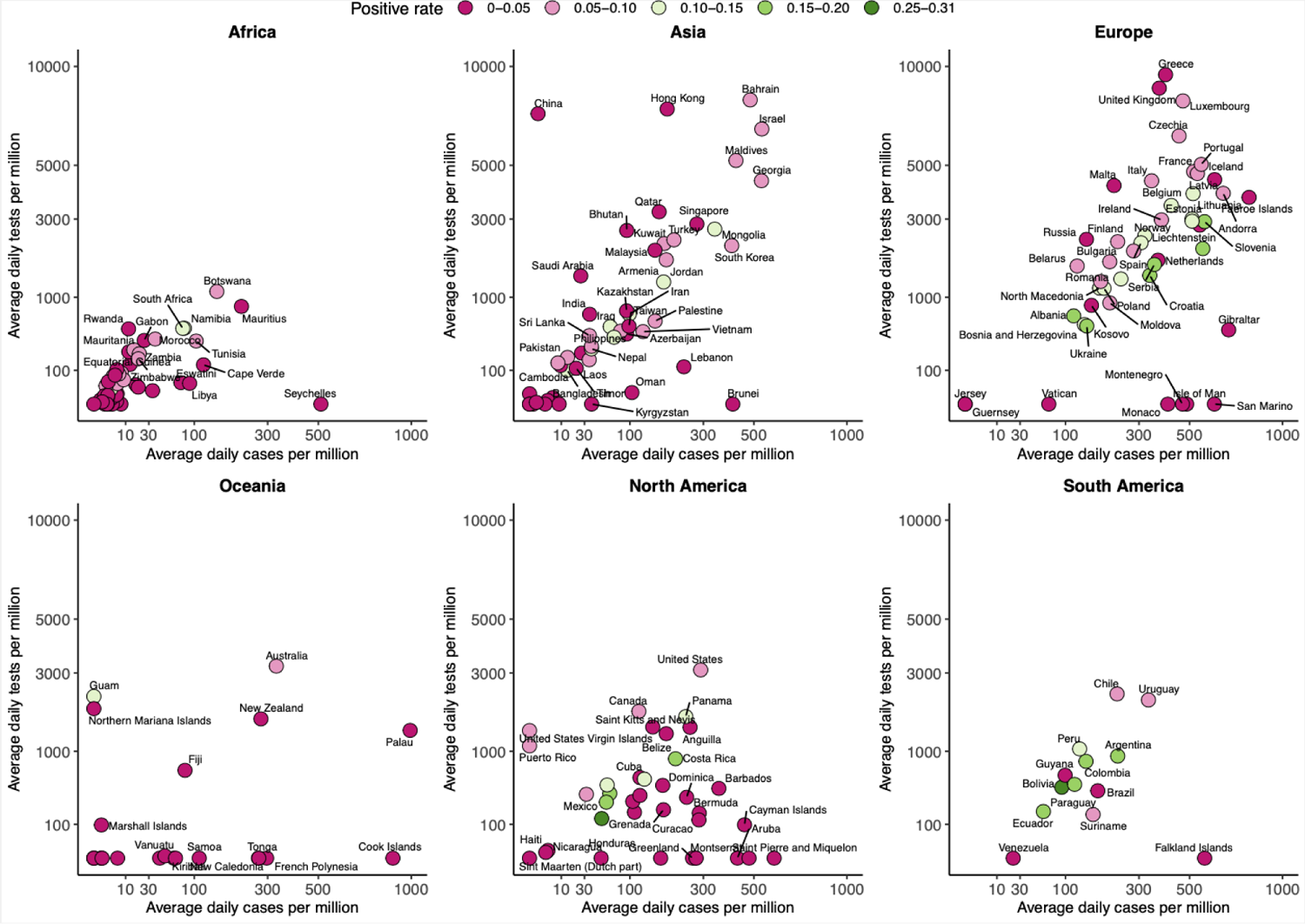
Global COVID-19 testing rates by continent. Graphs show testing rates per continent against recorded cases per million for countries with available data over the course of the pandemic. The extent of testing (average daily number of tests) is shown relative to the size of outbreaks (average daily number of reported cases) per million people per country. Data is obtained from Our World in Data (OWID). Colour scale denotes test positive rates. Points are shown for countries reporting the relevant data and with an average daily number of tests greater than zero for the relevant period. Axes are log scaled.

**Figure S9:**
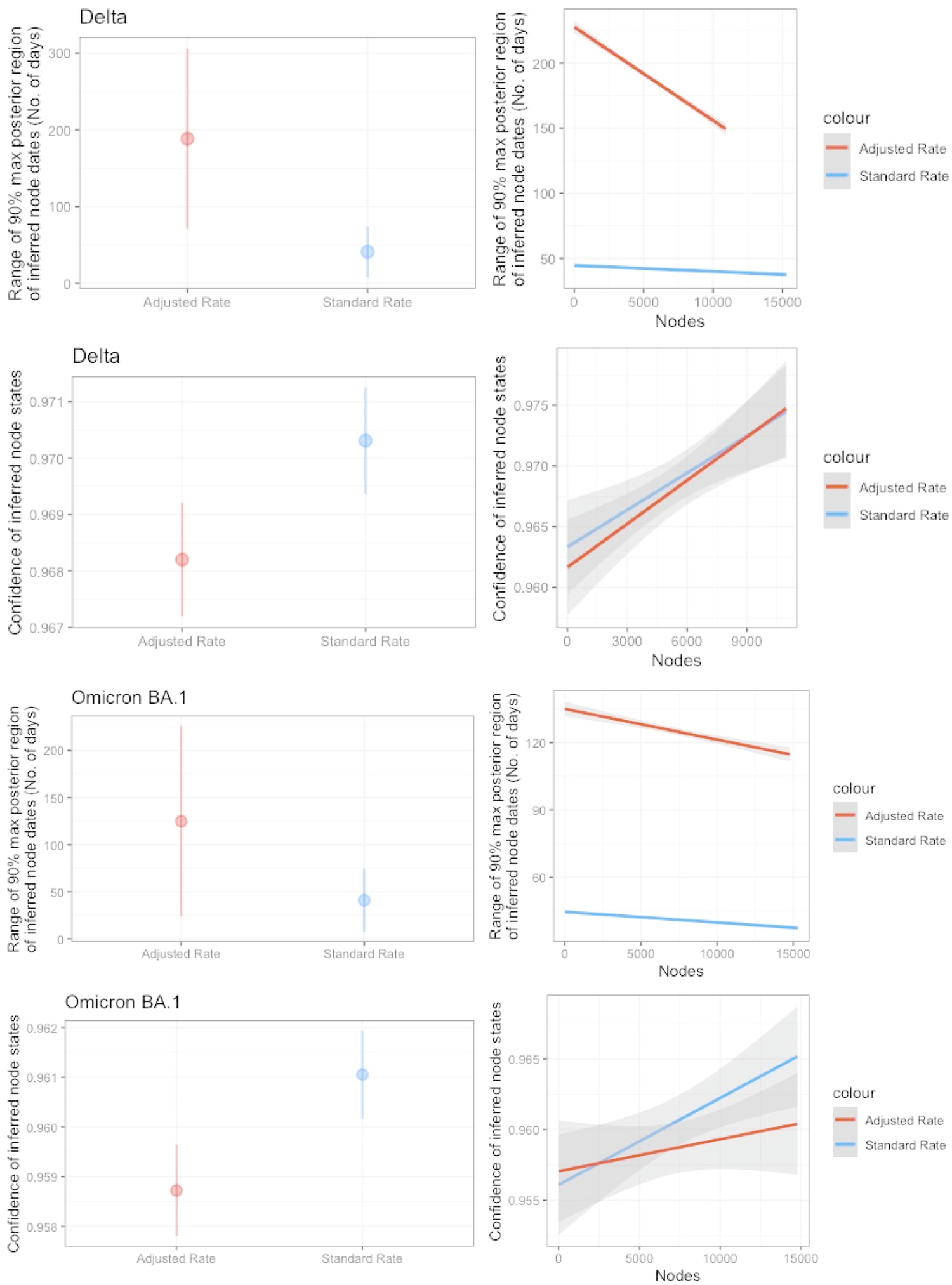
Justification for use of evolutionary rates. Graphs show the range of 90% maximum posterior region of inferred node dates (in number of days) and the confidence of reconstructed node states as proxies for robustness of inference either as an averaged measure for all nodes or by node number, from deepest nodes for the adjusted evolutionary rate v.s. the standard evolutionary rate. Results are shown for one phylogenetic reconstruction of Delta and Omicron BA.1 datasets.

**Table S1:** Published VOC tMRCA used in this study.

| **VOC** | **Method** | **Median** | **Upper** | **Lower** | **Source** |
| --- | --- | --- | --- | --- | --- |
| Alpha | BEAST | 2020-08-28 | 2020-09-09 | 2020-08-15 | Hill et al., 2022<br>(https://doi.org/10.1093/ve/veac080) |
| Beta | BEAST | 2020-08-05 | 2020-08-30 | 2020-07-15 | Tegally et al., 2021<br>(https://doi.org/10.1038/s41586-021-03402-9) |
| Gamma | BEAST | 2020-11-15 | 2020-11-24 | 2020-10-06 | Faria et al., 2021<br>(https://doi.org/10.1126/science.abh2644) |
| Delta | BEAST | 2020-10-19 | 2020-11-29 | 2020-09-06 | McCrone et al., 2022<br>(https://doi.org/10.1038/s41586-022-05200-3) |
| Omicron BA.1 | BEAST | 2021-10-09 | 2021-10-20 | 2021-09-30 | Viana et al., 2022<br>(https://doi.org/10.1038/s41586-022-04411-y) |
| Omicron BA.2 | BEAST | 2021-11-05 | 2021-11-29 | 2021-10-09 | Tegally et al., 2022<br>(https://doi.org/10.1038/s41591-022-01911-2) |
| Omicron BA.4/BA.5 | BEAST | 2021-12-15 | 2022-02-06 | 2021-11-25 | Tegally et al., 2022<br>(https://doi.org/10.1038/s41591-022-01911-2) |

